# Optimizing telework in an epidemic context: contrasting the infectious and non-communicable diseases perspectives

**DOI:** 10.1101/2024.07.18.24310632

**Authors:** Léo Moutet, Quentin J. Leclerc, Maylis Layan, Karim Aït Bouziad, William Dab, Paul Henriot, Elise Hodbert, Narimène Louati, Aurélie Maurin, Frédérique Thonon, Sylvie Znaty, Mohamed Ben Halima, Kévin Jean, Laura Temime

## Abstract

1.

**Objectives:** In a global context of increasing telework, this study explores its health effects, to determine if there is an optimal teleworking frequency during an epidemic. We aim to quantify the relationship between teleworking frequency and both infectious disease (ID) transmission and non-communicable disease (NCD) risk.

**Methods:** We developed a mathematical model simulating ID transmission and NCD acquisition in a medium-sized company, analysing how different teleworking levels impact workers health. We conducted a rapid literature review to identify potential exposure-response relationships between teleworking and NCD risk and inform this model. We then simulated infection dynamics over a three-month epidemic wave to contrast ID and NCD risks in relation to the extent of telework engagement.

**Results:** Evidence from the literature showed varying patterns of NCD risk across different teleworking frequencies. Depending on these relationships, we observed that risk may peak at low, intermediate, or high teleworking levels. We demonstrated the existence of a benefit-risk balance between reducing ID transmission and potentially increasing NCD burden.

**Conclusions:** Based on current scientific knowledge, no single optimal teleworking frequency can be identified, as the optimum varies depending on the NCD outcome considered. Our study highlights the need for stronger evidence to estimate robust exposure-response functions linking teleworking frequency and NCDs, and ultimately to inform prevention strategies for both infectious and NCD risks in an epidemic context.

## 2. Introduction

Teleworking describes work that is fully or partially carried out at a location other than the default place of work, typically at home (1). In the European Union, the proportion of workers who sometimes or usually telework rose from 9% in 2019 before the COVID-19 pandemic, to 12.2% in 2022 (15.8% to 21.3% in France) (2). Recently, several large organisations have switched to a full-remote work organisation, where workers whose job can be performed remotely no longer have an office (3,4). During the same period, the willingness of employees to telework has increased (5,6). Overall, the increasing prevalence of teleworking implies the necessity to understand the impact of this activity on worker health (7).

From the perspective of infectious diseases (IDs), there is a consensus that teleworking in the context of an epidemic reduces the risk of infection (8,9). An important proportion of contacts occur at work, hence reducing these can have a substantial impact on ID incidence (10–14). The value of teleworking against ID transmission was particularly highlighted during the COVID-19 pandemic, during which public health and social measures led to a global transition towards telework across numerous sectors (15). Through these elements, we can establish an exposure-response relationship between teleworking and IDs, whereby an increased teleworking frequency mechanistically leads to a reduction of ID risk.

On the other hand, the shape of this potential exposure-response relationship between teleworking and non-communicable diseases (NCDs) risk remains unclear (16,17). This is notably due to conflicting evidence for the impact of teleworking on NCD risk, within which we can distinguish between physical and mental health. In a well-organised manner, teleworking can enhance the equilibrium between work-related commitments and personal life, diminish road congestion and commuting time, and decrease atmospheric pollutants, all of which indirectly contribute to the improvement of physical and mental well-being (18,19). However, suboptimal physical settings and workplace design can cause musculoskeletal disorders (MSDs), ocular strain (20), and an associated psychological burden (21). Other major health determinants can be affected by teleworking such as obesity, alcohol abuse, physical inactivity, and tobacco use (22). Combined, these elements suggest that the relationship between teleworking and NCD risk is not as clear as for IDs. By extension, this implies that the overall impact of teleworking on health and the potential existence of an optimal teleworking frequency to maximise health benefits are unclear. In this study, we define as “optimal” a teleworking frequency that would yield the lowest possible number of COVID-19 cases, and the lowest possible number of NCD cases.

In this study, we aimed to explore the health impacts associated with telework in an epidemic context, including both ID and NCD. We designed a novel mathematical model simulating the incidence of an ID amongst employees of a non-specific company. This model accounts for the impact of teleworking frequency on both the ID transmission and the incidence of NCD. We parameterised the model by conducting a rapid review to identify exposure-response relationships that have been quantified between teleworking frequency and physical or mental health. We then illustrated how different exposure-response functions may lead to contrasting impacts of teleworking on health.

## 3. Methods

### 3.1. Mathematical model

To simultaneously capture the relationship between teleworking and both infectious disease (ID) and non-communicable disease (NCD) risk, we designed a compartmental deterministic model to represent the population of employees from a non-specific company during an epidemic wave of a SARS-CoV-2-like virus (Figure 1a). This model is summarised in the following set of ordinary differential equations driving the dynamics of numbers of individuals in each model compartment:

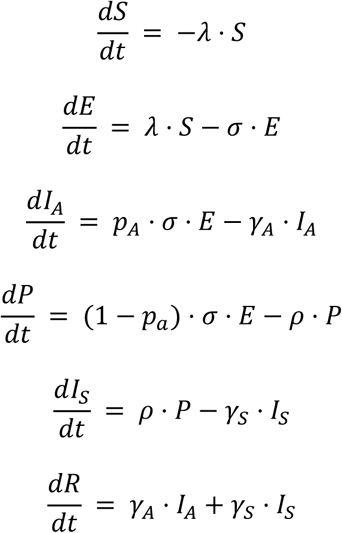

**Figure 1:**
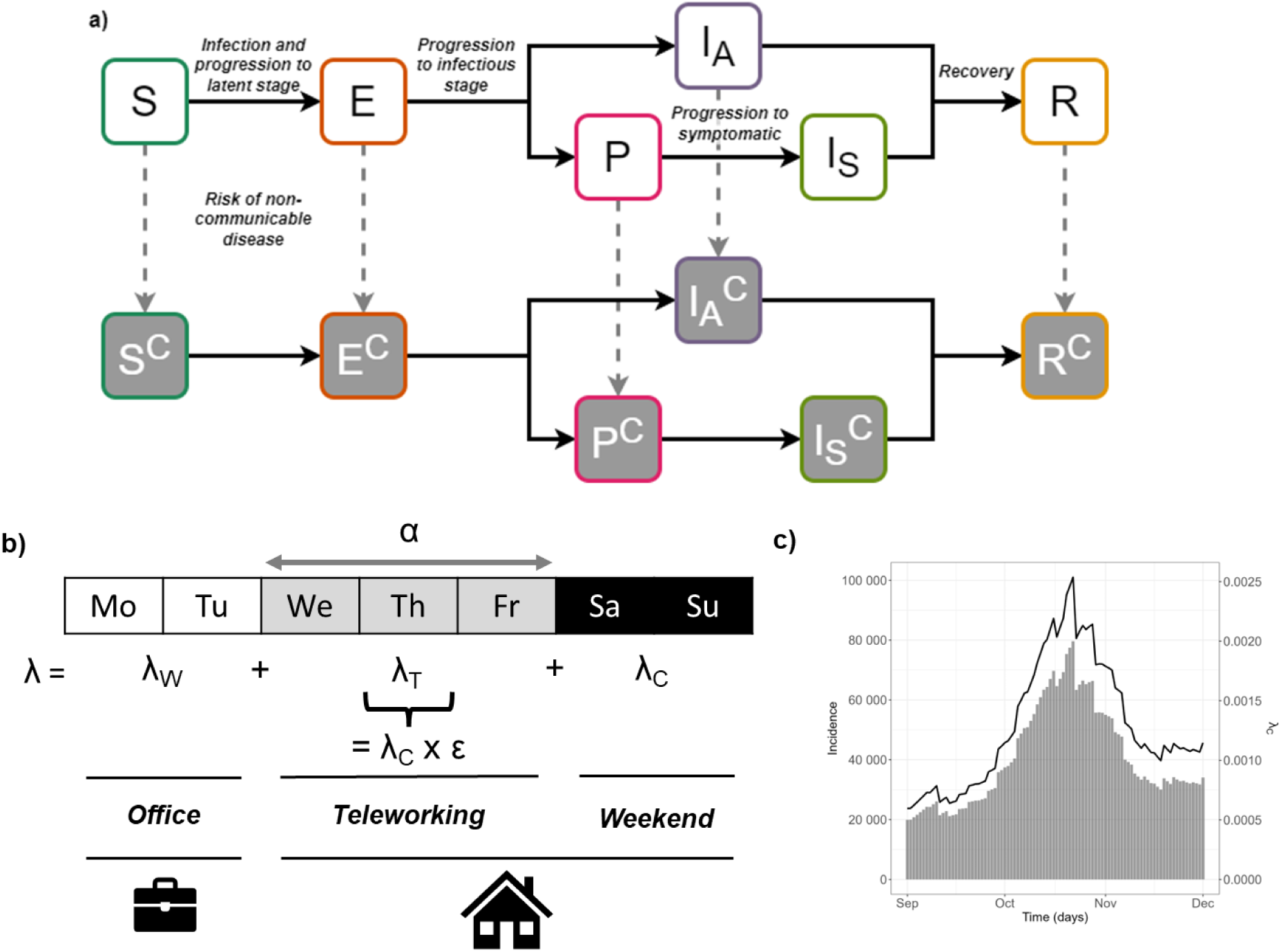
Model structure. a) Model diagram. With regards to the infectious disease, individuals can either be Susceptible (S), Exposed (E), Infected Asymptomatic (I_A_), Presymptomatic (P), Infected Symptomatic (I_S_) or Recovered (R). Compartments with the superscript “C” indicate individuals who will eventually develop a NCD due to teleworking. **b) Components of the total force of infection *λ*. c) Community epidemic curve (bars, left axis) and corresponding force of infection *λ_c_* (line, right axis).**

We considered that all employees of the company are initially susceptible (*S*) to the ID and can become exposed (*E*), *i.e.* infected but not yet infectious, following a transmission event at rate *λ*. Exposed individuals progress through a latent period at rate *σ* before either becoming infectious asymptomatic (*I_A_*, with probability *p_A_*), or infectious presymptomatic (*P*). Presymptomatic individuals eventually become infectious symptomatic (*I_S_*) at a rate *ρ*. All infectious individuals eventually become recovered (*R*) at a rate which varies depending on whether they are asymptomatic (*γ_A_*) or symptomatic (*γ_S_*). The total number of employees N=S+E+I_A_+P+I_s_+R is assumed constant over the time period of interest. The cumulative incidence is calculated over a 3-month period.

#### 3.1.1. Rate of infection

Employees can become infected either via contacts with 1) other infectious employees during working days when they are present in the workplace, 2) contacts with individuals outside the company during working days when they are teleworking, or 3) contacts with individuals outside the company during non-working days (Figure 1b). We assumed that five days per week are working days, and calculated the overall force of infection as a weighted average of forces of infection in the workplace (*λ_W_*), during telework (*λ_T_*), and on non-working days (*λ_C_*), as follows:

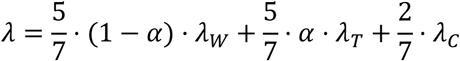

with

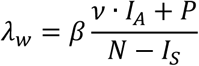

Here, *α* indicates the average proportion of time spent in telework rather than in the office. If employees are working from the office, they can be infected at a rate *β* either by the proportion of infected asymptomatic (*I_A_*) or presymptomatic (*P*) employees also present. We assumed that infected symptomatic (*I_S_*) individuals are systematically on sick leave, and hence cannot infect other employees since they are absent from the workplace. We further assumed that asymptomatic individuals are less infectious than presymptomatic by a factor *ν* (23). The term *λ_C_* corresponds to the force of infection which an employee is subjected to outside the workplace, on a non-working day. We considered that this represents the wider epidemic in the community, which is independent of the workplace epidemic since it is much larger. We calibrated *λ_C_* to available incidence data from the second wave of SARS-CoV-2 in France (01/09/2020 - 01/12/2020) (Figure 1c). On teleworking days, we assumed that employees would also have contacts with other individuals in the community, although at a lower rate than on non-working days, hence we defined the force of infection on teleworking days *λ_T_* as a fraction *ε* of *λ_C_*.

We calculated the within-company basic reproduction number *R_0_* using the next generation matrix method (24) (Supplementary Text 1 (25)) from which we derived the per-employee transmission rate *β* for presymptomatic individuals in the workplace as

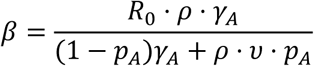

#### 3.1.2. Rate of non-communicable disease incidence

To represent the incidence of the selected NCD due to telework in this model, we duplicated the compartments listed above to further stratify individuals according to whether they will develop a NCD due to teleworking. We considered that individuals in the infected symptomatic state cannot move to the corresponding compartment, since they do not telework but are on sick leave. For all other states, the transition towards the NCD status occurs at an average daily rate defined by the exposure-response function *f(α)* which depends on the proportion of telework *α*. Possible shapes for this function were informed by the studies identified in our rapid review (22,26,27). We derived the exposure-response curves using linear interpolation between the data points retrieved from the identified studies. To ensure comparability between the different forms of *f(α)*, we systematically set the baseline value *f*(*0*) equal to 8.9 x 10^-5^ per day. This corresponds to an annual probability of approximately 3.2% (= 1 – exp(-8.9*10^-5^*365)) to develop a NCD in absence of teleworking, similar to the Global Burden of Disease (GBD) 2021 estimates for the cumulative incidence of MSDs and mental disorders (28). Parameter values for the model are summarised in Table 1.

**Table 1:** Model parameters.

| Name | Symbol | Value | Range explored | Source |
| --- | --- | --- | --- | --- |
| Reproduction number | $R_0$ | 2.66 | 2 - 4 | (29) |
| Coefficient of relative infectiousness of asymptomatics | $\nu$ | 0.35 | 0.1 – 1.27 | (23) |
| Coefficient of relative community force of infection on teleworking days | $\varepsilon$ | 0.21 | 0.17 – 0.25 (assumed +/- 20%) | (15) |
| Progression rate from exposed to infectious (= 1/incubation period) | $\sigma$ | $1/6.57 \text{ days}^{-1}$ | $1/18.87 - 1/1.80 \text{ days}^{-1}$ | (30) |
| Probability of asymptomatic infection | $p_A$ | 0.2 | 0.17 – 0.25 | (23) |
| Recovery rate from infection for asymptomatics | $\gamma_A$ | $1/5 \text{ days}^{-1}$ | N/A | (31) |
| **Progression rate from presymptomatic to symptomatic** | $\rho$ | $1/1.5 \text{ days}^{-1}$ | N/A | (31) |
| **Recovery rate for symptomatics** | $\gamma_s$ | $1/5 \text{ days}^{-1}$ | N/A | (31) |
| **Baseline incidence rate of non-communicable disease in absence of telework** | $f(0)$ | $8.9 \times 10^{-5} \text{ days}^{-1}$ | N/A | (28) |
| **Maximum change in relative risk of non-communicable disease due to telework\*** | $\omega$ | +/- 0.7 | +/- 0.05-0.7 | This study |

\*This parameter is only used in the sensitivity analysis

Model simulations over a 3-month time period were performed using R version 4.2.2. The code for the model and all the analysis presented in this article is freely available online (32).

#### 3.1.3. Converting incidences to health burden

In order to compare the health burden of ID and NCD cases using the same metric, we estimated the number of disability-adjusted life years (DALYs) caused by these diseases, placing ourselves in a scenario where the ID was COVID-19 and the NCD was lower back pain. For COVID-19, DALYs were computed by adding up the years of life lost (YLL) from deceased individuals and the years of healthy life lost due to disability (YLD) from symptomatic cases, while the DALYs for the NCD were simply the YLDs. Disease-specific disability weights were obtained from the 2021 GBD (28). In the GBD, these weights are stratified by the level of severity of the disease. In order to obtain disease-specific average disability weights which account for the severity spectrum of each disease, we divided disease-specific YLD-estimates by the number of prevalent cases among individuals aged 20 to 64 years in France, as provided by the GBD. We also derived average disease durations from the 2021 GBD estimates using the ratio of prevalent to incident cases. For both diseases, we obtained the YLD term by multiplying the model-predicted number of cases by the average disease-specific disability weight, and the average disease duration. Finally, we estimated the YLLs from deaths following SARS-CoV-2 infections by multiplying the model-predicted number of COVID-19 cases by the age-specific infection fatality rate (33) and the age-specific life expectancy. Based on the statistics of the French public finance services on the working population, the mean age was assumed to be 41 years (https://www.insee.fr/fr/outil-interactif/6794598/EVDA/FRANCE). In 2023, life expectancy at 41 was 45.6 years for women and 40.5 for men, averaging to 43.1 years.

### 3.2. Rapid review

We conducted a rapid review of research articles investigating quantifiable aspects of telework (e.g. frequency measured in days per month or hours per week) in association with the risk of NCDs, such as mental health disorders and MSDs (34).

#### 3.2.1. Search strategy and study selection

Previous reviews on the impact of telework on health have found that the most notable outcomes are related to behavioural risks, mental health or MSDs (35–37). Therefore, we restricted our review to the impact of telework on these outcomes.

We searched three databases (Pubmed, Scopus and Google Scholar) to identify original studies assessing the quantitative association between telework and health. We then synthesised the evidence regarding the shape and magnitude of the relationship between telework exposure and the various health outcomes studied. The search terms used were (“telework” OR “remote work” OR “work from home” OR “telecommuting”) AND (“mental health” OR “musculoskeletal disorder”).

#### 3.2.2. Inclusion and exclusion criteria

We used the following inclusion and exclusion criteria to select studies:

- Publication type: we included original research articles only.
- Population: we included articles studying workers exposed to some level of telework, whether they were part of a specific sub-population (such as specific professions) or not.
- Exposure: we included studies that compared at least two levels of exposure to telework, in addition to no telework.
- Outcomes: we included studies that reported MSDs, mental health conditions or behavioural risks as outcomes, and excluded studies reporting only other outcomes. We did not restrict ourselves to a single specific MSD, mental health condition or behavioural risk.

We placed no restriction on the date of publication or the language.

#### 3.2.3. Data collection

From the selected studies, we extracted the following information: study years, country, working population (sector), study design, quantification of telework exposure, the health-related outcomes examined, and the shape of the observed relationship. Importantly, we extracted relationships regarding health-related outcomes for any telework level different from 0 to portray plausible exposure-response functions. To inform the model, we only considered exposure-response relationships based on significant (p < 0.05) values.

### 3.3. Sensitivity analysis

#### 3.3.1. Theoretical exposure-response functions

To compare the exposure-response relationship shapes independently of the upper/lower bounds of the functions, we developed a set of theoretical functions which reproduced the shapes we identified in our review, whilst allowing us to flexibly change the upper/lower bounds. These functions all take as input the teleworking frequency α (α ∈ {0,1}) and return the relative risk of non-communicable disease, with the constraint that f(0) = 1 and the upper/lower bound is equal to ω.

For linear increasing or decreasing functions:

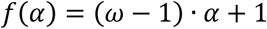

Where the function is linear increasing if ω > 1 or decreasing if ω < 1.

For inverted-U or U-shaped functions:

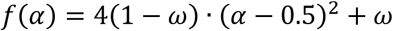

Where the function is inverted-U shaped if ω > 1 or U-shaped if ω < 1. The value ω is reached for α=0.5.

For L-shaped functions:

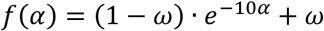

The choice of value 10 controls the rate of decay of the function, to obtain a function similar to the observed LS function from the literature with ω=0.3 represented in Supplementary Figure 1.

These theoretical functions are represented in Supplementary Figure 2.

#### 3.3.2. Impact of model parameter values on incidence

We calculated partial correlation coefficients to examine the impact of model parameter values on cumulative incidence of ID and NCD over the period, assuming a teleworking frequency of 0.5. A total of 400 parameter sets were drawn, with each parameter independently sampled from a uniform distribution within the ranges specified in Table 1.

## 4. Results

### 4.1. Studies on NCD and telework identified to inform the model

From our rapid review, we identified only three studies that met our selection criteria and presented exposure-response relationships between teleworking and NCD risk which could inform the mathematical model (Table 2). Of those studies, one focused on work-related outcomes (26), one on physical health (27), and one on health determinants (including behavioural changes) impacting both mental and physical health (22). Importantly, two of the identified studies were entirely conducted before the COVID-19 pandemic (22,26).

**Table 2:**
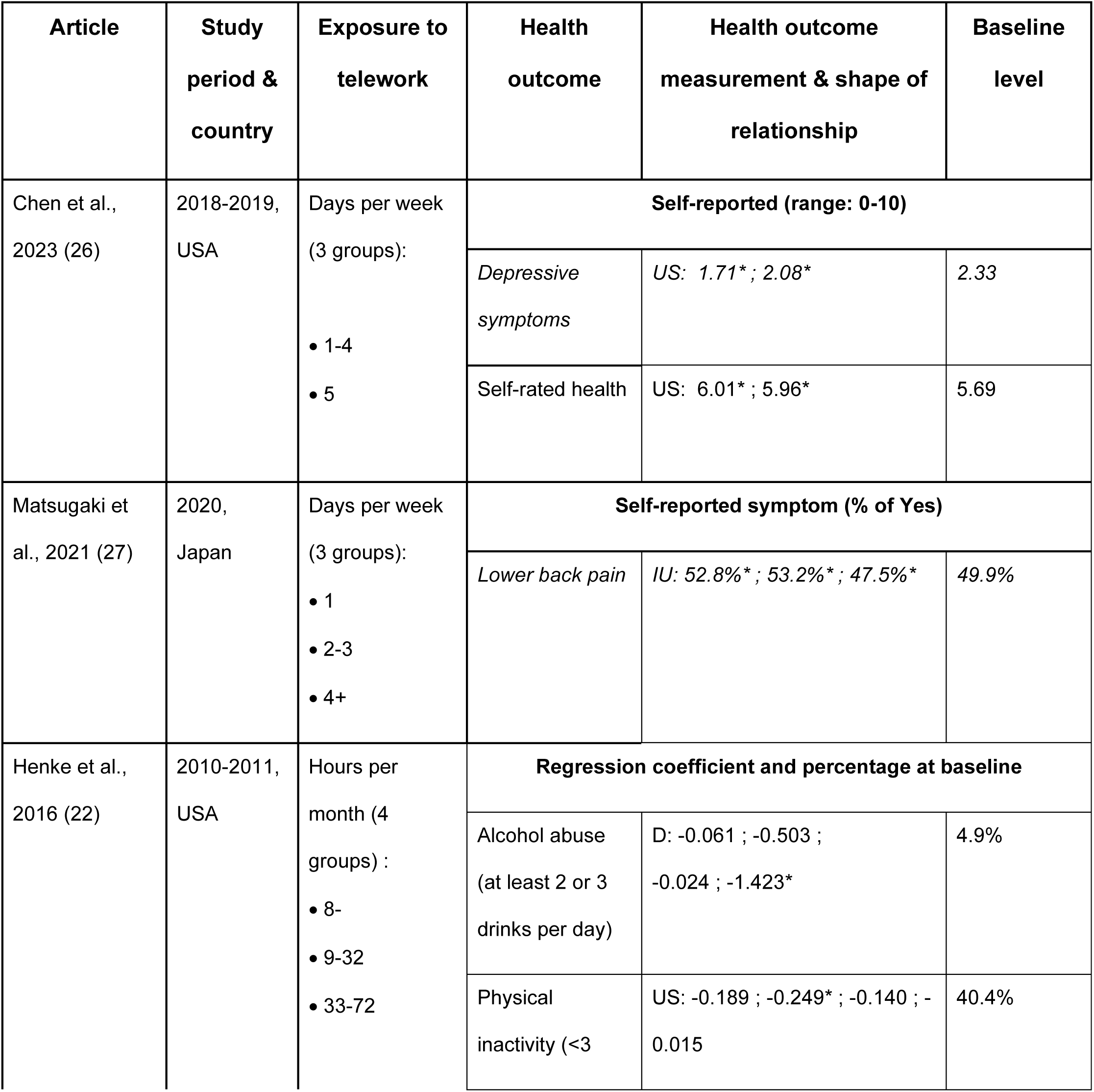
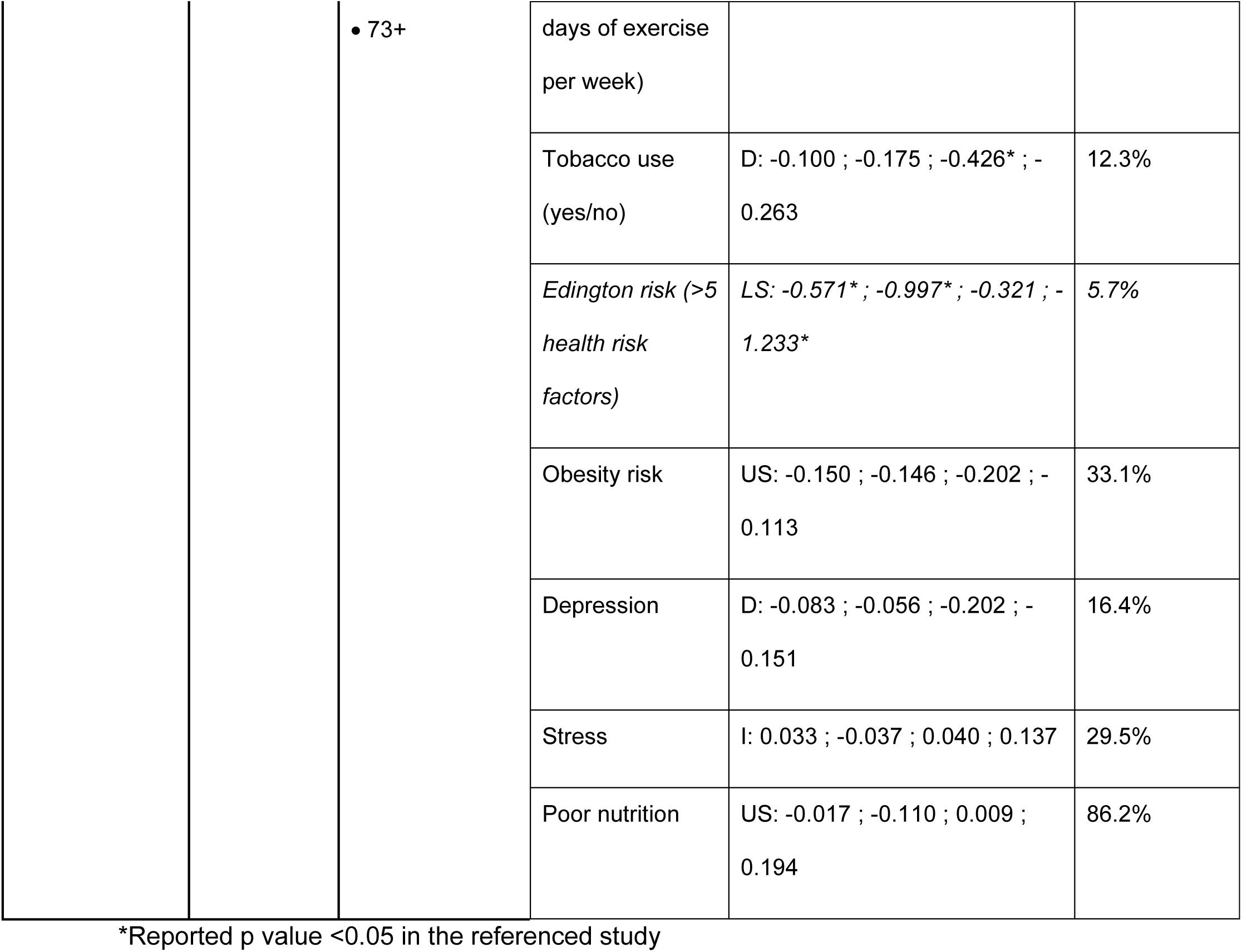
Summary of identified studies on the exposure-response relationship between teleworking frequency and non-communicable disease (NCD) risk. Here, we classified (or retrieved) the shape exposure-response relationships into five types: U-shaped (US), inverted U-shaped (IU), L-shaped (LS), broadly decreasing (D) and broadly increasing (I). Relationships in italic are tested in our mathematical model in the next section. “Baseline level” values are values from the studies for groups not exposed to telework (0 days per week).

The study that focused on work-related outcomes notably examined depressive symptoms and self-rated health (26). The results of this longitudinal study conducted on United States employees highlighted exposure-response relationships for several outcomes. A U-shaped (US) relationship was found between the number of teleworking days per week and depressive symptoms, with a higher risk among non-teleworkers (0 days per week) and those with a higher frequency of telework (5 days per week). The association between number of teleworking days per week and self-rated health also displayed a US relationship.

For physical health, the study we identified only focused on lower back pain as a MSD-related outcome (27). The findings from this cross-sectional study conducted in Japan revealed an inverted U-shaped (IU) exposure-response relationship between teleworking frequency and lower back pain, with the highest risk in case of intermediate teleworking frequencies (2-3 days/week).

Finally, the last study described exposures related to both mental and physical health such as alcohol abuse or physical inactivity (22). This longitudinal study was conducted in the United States on insurance company employees. Retrieved results (significant values at p < 0.05) suggested a broadly decreasing relationship between teleworking intensity and risks of alcohol abuse and tobacco use. For the Edington risk score (a summary score accounting for several risk factors), we observed an L-shaped (LS) relationship with increased telework intensity, with an initial rapid decrease in risk at low telework frequencies followed by a more stable relationship at higher frequencies.

### 4.2. Impact of varying teleworking frequency on health outcomes

Our review shows that the exposure-response relationship between teleworking and NCD risk may vary depending on the mental or physical health focus, as well as on other factors. This is bound to impact the level at which teleworking best prevents both infectious disease (ID) and NCD. Based on our literature review and the statistically significant relationships we identified, we explored three relationships between telework frequency and the risk of NCD in our model: L-shaped (LS, for Edington risk), U-shaped (US, for depressive symptoms), and inverted U-shaped (IU, for lower back pain). In the model, the corresponding exposure-response functions *f(α)* giving the daily rate at which individuals will develop a NCD for a given frequency of teleworking *α* were parameterized using data from the three studies analysed in the rapid review (22,26,27) (values in italic in Table 2, see Supplementary Text 2 for details). Importantly, since we used values from the literature to parameterize these functions, the strengths of the association and therefore their upper and lower bounds are different.

Implementing telework throughout the epidemic wave substantially reduced the cumulative incidence of ID amongst employees (yellow curve), with greater reductions at higher teleworking frequencies and no substantial differences between frequencies greater than 0.8, i.e. 4/5 days per week (Figure 2a). However, the cumulative incidence of NCD at different teleworking frequencies varied depending on the assumed exposure-response relationship function, with the predicted peak incidence of NCD occurring either at low (LS, US) or intermediate (IU) teleworking frequencies (Figure 2b).

**Figure 2:**
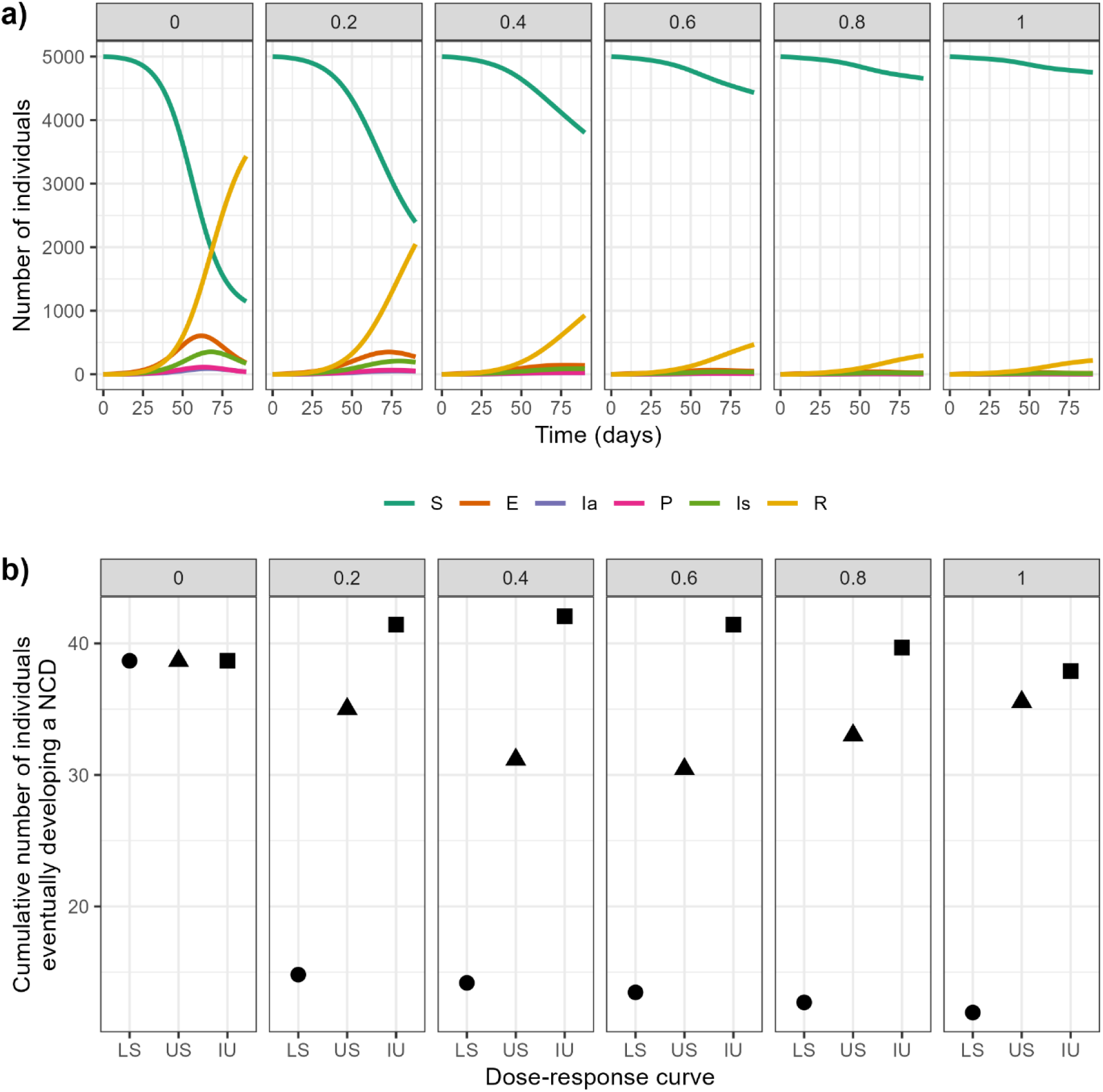
Epidemic dynamics and cumulative incidence of non-communicable disease (NCD) at varying telework frequencies. Model simulations over 3 months using a telework frequency *α* = 0 (no telework), 0.2 (1 day in 5), 0.4 (2 days), 0.6 (3 days), 0.8 (4 days) and 1 (full telework), for **a)** changes in the number of susceptible (S), exposed (E), asymptomatically infected (Ia), presymptomatic (P), symptomatically infected (Is) and recovered (R) employees, and **b)** the cumulative incidence of employees who will eventually develop a NCD, in case of a L-shaped (LS, circles), U-shaped (US, triangles) or inverted U-shaped (IU, squares) exposure-response relationship between telework and NCD risk.

### 4.3. Impact of varying teleworking start date on health outcomes

The impact of teleworking on cumulative disease incidence further varied depending on the timing of its implementation from the start of the epidemic wave (Figure 3). Since we performed simulations in the context of an epidemic wave, implementing teleworking too late prevented any significant reduction of the ID cumulative incidence (orange lines). If we set an arbitrary objective to reduce this incidence below 50% of the baseline value obtained without teleworking, at least two days of teleworking (frequency of 0.4) were necessary to reach this target, with teleworking implemented within the first 40 days of the epidemic (solid orange lines).

**Figure 3:**
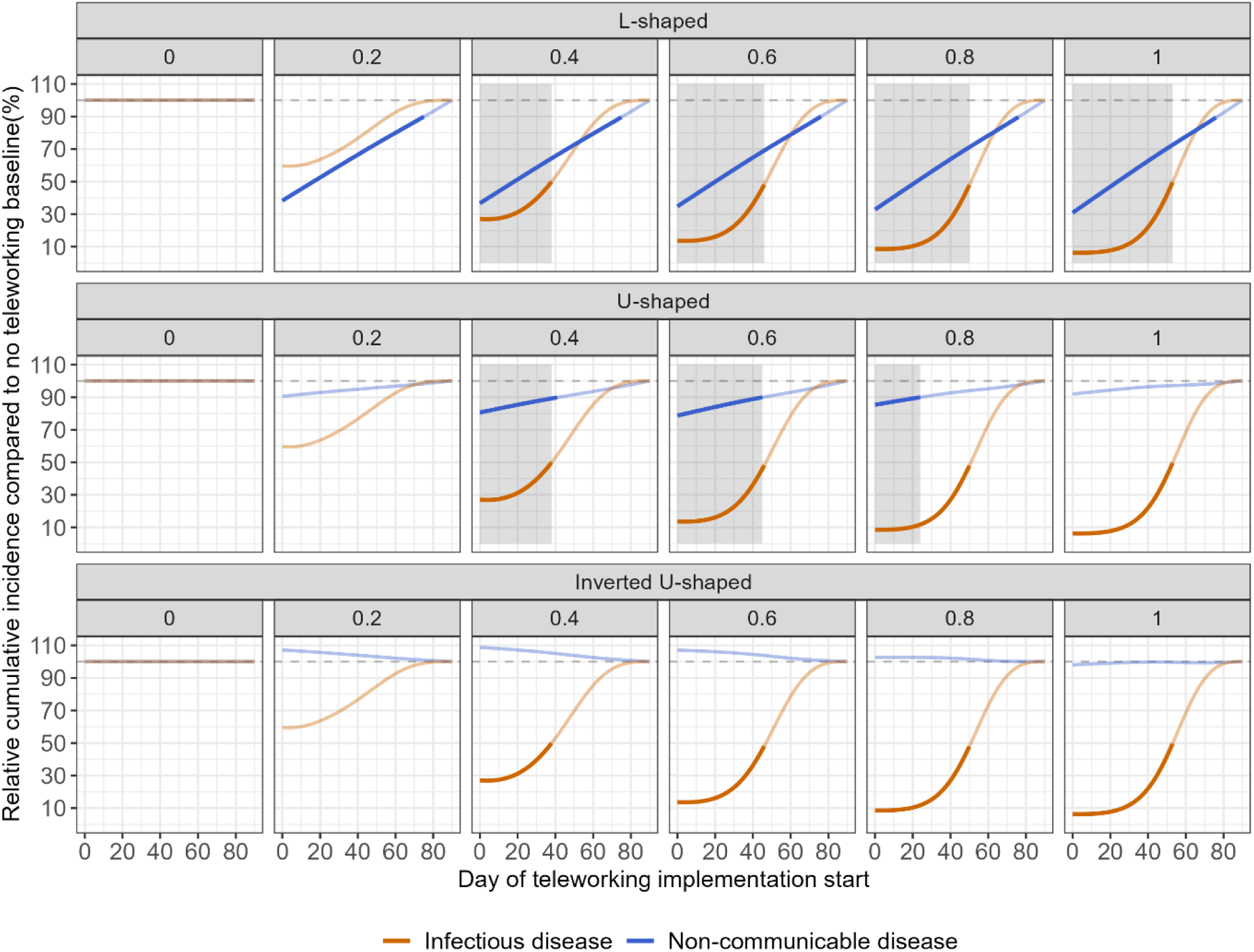
Impact of teleworking timing and frequency on the relative cumulative incidences of infectious (ID) and non-communicable diseases (NCD) using observed exposure-response functions. Cumulative incidence corresponds to the number of employees infected over the three months of the simulated epidemic. Here, we represent the cumulative incidences of ID and NCD relative to the cumulative incidences predicted by each model without teleworking (first column). For example, the lowest incidence of NCD (blue line) for the L-shaped function (first row) is when teleworking is implemented with a frequency of 1 (5 days a week) on day 0 (last column), while for the U-shaped exposure-response function (second row) this occurs at a frequency of 0.6 (fourth column). The grey dashed line indicates 100%, i.e. the baseline incidences. Lines are solid when the relative incidences of ID and NCD are respectively lower than 50% and 90% (chosen arbitrarily as examples), and faded otherwise. The shaded grey areas indicate conditions where both relative cumulative incidences are below the defined targets of 50% and 90% (for ID and NCD incidence, respectively).

Regarding the change in NCD risk caused by teleworking, as expected, early implementation of teleworking systematically led to a higher change in cumulative incidence (blue lines). However, the nature of this change depended on the assumed exposure-response function shape, upper and lower bounds, and on the teleworking frequency. For example, in the case of the L-shaped relationship for Edington risk parameterised according to our review (first row), the cumulative incidence of NCD was lowest when teleworking was implemented at a frequency of 1 on the first day (last column), while for the U-shaped relationship for depressive symptoms (second row) the lowest incidence was achieved at a frequency of 0.6 (fourth column). On the other hand, teleworking led to an increase in cumulative NCD incidence (relative incidence > 100%) when considering the inverted U-shaped relationship with lower back pain (last row), particularly at intermediate teleworking frequencies (0.4-0.6, third and fourth columns). In that case, slightly delaying the implementation of teleworking to avoid increasing the risk of NCD while still having an impact on ID could be better, and/or implementing 100% teleworking, which would not increase the risk of NCD while keeping the ID risk at a minimum. For the US relationship, timing is also important because early teleworking implementation for intermediate frequencies reduces both health risks below their respective targets, which is not the case for lower or higher frequencies. Additionally, since there is no major difference for ID relative risk between frequencies of 0.8 and 1, a teleworking frequency of 0.8 would be preferable to achieve strong reductions in both NCD and ID risks.

The contrasting relationship between teleworking and ID versus NCD risks implies that reducing both incidences simultaneously may not always be feasible, depending on the characteristics of the NCD. In our illustrative example here, where we aimed to reduce ID incidence by at least 50% and NCD incidence by at least 10%, we observed only limited conditions where teleworking could simultaneously reduce both diseases incidences below these targets (Figure 3, grey shaded areas). The definition of an “optimal” teleworking frequency to improve health is therefore inconsistent and will vary depending on (*i*) the observed exposure-response relationship between teleworking and health outcomes, and (*ii*) the relative importance granted to both ID and NCD risks when defining target thresholds.

### 4.4. Exploring different exposure-response relationships between teleworking and NCDs

While the exposure-response functions used in the previous section were directly informed by values we identified in the literature, their upper and lower bounds varied, which introduces a bias when comparing the incidence of different NCDs since the differences are not only attributable to the shape of the functions. For example, the greatest change in relative risk of NCD for the IU function (lower back pain) was approximately +6%, while for the LS function (Edington risk) this reached -70%. To overcome heterogeneity in exposure categorisation and data limitations regarding exposure-response curves, we reproduced our analysis using five theoretical functions with shapes corresponding to those we identified in our review (see Supplementary Text 2 for details, and Supplementary Figures 2 and 3 for the shapes of the functions), but each with the same greatest absolute change in relative risk of NCD (+/- 70%). The results obtained underline how the shape of these functions alone affects the optimal telework frequency and implementation date, independently of upper/lower bounds (Figure 4). This analysis also highlights scenarios where the relative increase in NCD incidence can be equivalent to the relative decrease in ID incidence, as can be seen for the theoretical IU function at a telework frequency of 0.6, and the theoretical linear increasing function at a frequency of 1 (Figure 4, rows 3 and 4).

**Figure 4:**
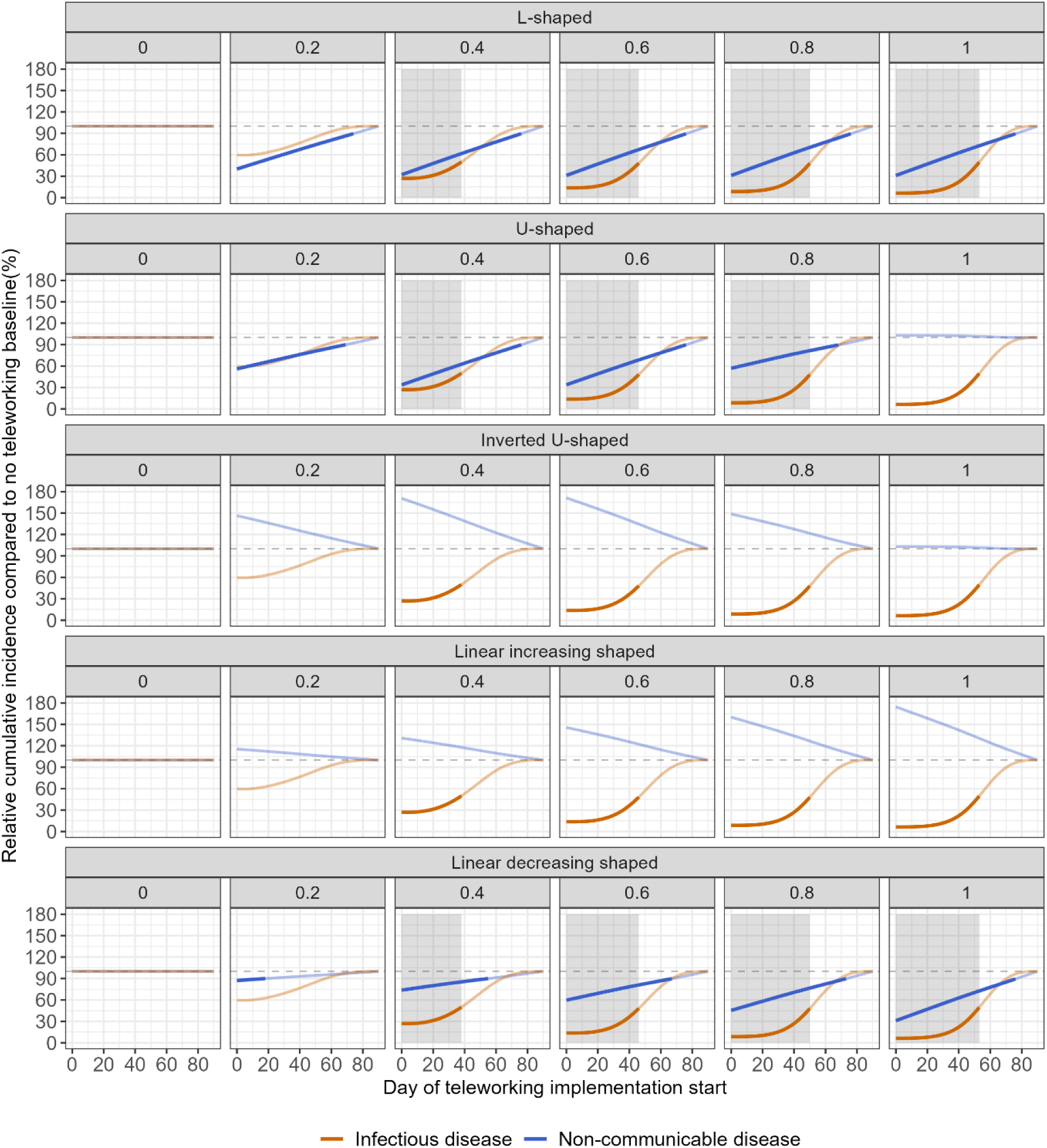
Impact of teleworking timing and frequency on the relative cumulative incidences of infectious (ID) and non-communicable diseases (NCD) using theoretical exposure-response functions. Cumulative incidence corresponds to the number of employees infected over the three months of the simulated epidemic. Here, we represent the cumulative incidences of ID and NCD relative to the cumulative incidences predicted by each model without teleworking (first column). The grey dashed line indicates 100%, i.e. the baseline incidences. Lines are solid when the relative incidences of ID and NCD are respectively lower than 50% and 90% (chosen arbitrarily as examples), and faded otherwise. The shaded grey areas indicate conditions where both relative cumulative incidences are below the defined targets of 50% and 90% (for ID and NCD incidence, respectively).

### 4.5. Estimating the health burden of infectious and non-communicable diseases at different teleworking frequencies

At baseline, without any telework, we estimate that an infectious disease such as COVID-19 could be responsible for approximately 257 DALYs in our worker population (96% of total health burden, Figure 5). In the same scenario, a non-communicable disease such as lower back pain could be responsible for approximately 11 DALYs (4% of total burden).

**Figure 5:**
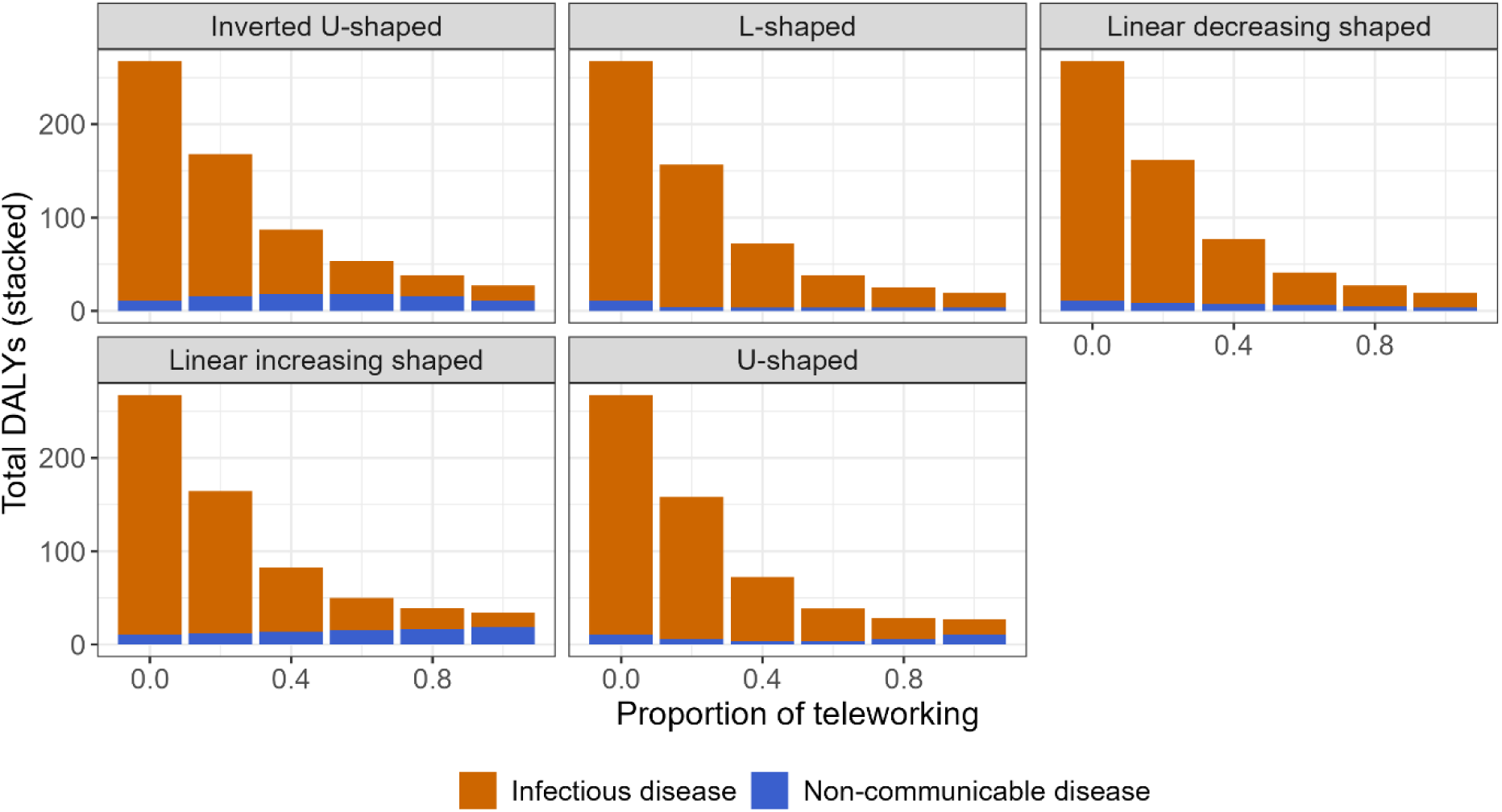
Impact of teleworking frequency on the health burden of infectious (ID) and non-communicable diseases (NCD) using theoretical exposure-response functions. The burden is expressed in disability-adjusted life years (DALYs). The health burden is estimated using COVID-19 as the infectious disease, and lower back pain as the non-communicable disease, in a population of 5 000 workers with an average age of 41 years.

As teleworking frequency increases, the burden of the ID decreases, down to 16 DALYs at a frequency of 1. However, depending on the exposure-response function, the burden of the NCD can either increase or decrease, while its value relative to the burden of the ID always increases. For example, assuming an L-shaped or linear decreasing shaped function, the NCD burden remains small, but represents a higher share of total health burden (3 DALYs, 15% of total burden at 100% teleworking frequencies). However, with a linear increasing shaped function, the burden of the NCD can increase up to 18 DALYs, representing 53% of the total health burden of the ID and NCD combined.

### 4.6. Sensitivity analysis

The only differences in correlation coefficients that we observed between scenarios of exposure-response relationships affected one parameter, the maximum change in relative risk of NCD due to telework (*ω*) (Supplementary Figure 3); as expected, this parameter was strongly positively correlated (coefficient ≈ 1) with NCD incidence for IU and LI curves, and strongly negatively correlated (coefficient ≈ -1) with NCD incidence for LD, LS and US curves.

Regardless of which exposure-response function was used, the relative infectiousness of asymptomatic individuals (*ν*) was only slightly negatively correlated with ID cumulative incidences (coefficient ≈ 0.2). The coefficient of relative community force of infection on teleworking days (*ε*) was only slightly positively correlated with ID incidence (coefficient ≈ 0.2).

The reproduction number (R_0_) and the progression rate from exposed to infectious (*σ*) were strongly positively correlated with ID incidence (coefficient > 0.7). These two parameters were negatively correlated with NCD incidence (coefficient ≤ -0.5), which is expected since we assume that infected individuals cannot develop a NCD during the period when they are infected and symptomatic. Inversely, the proportion of asymptomatic infections (*p_A_*) was slightly positively correlated with NCD incidence (coefficient ≈ 0.2), since it leaves a greater proportion of infected individuals at risk of developing a NCD in parallel due to reduced sick leaves.

## 5. Discussion

Recent years have seen an unprecedented increase in teleworking frequency in many countries worldwide. In this study, we aimed to determine if there existed an optimal teleworking frequency to maximise health benefits, by designing a mathematical model accounting for both infectious disease (ID) transmission and NCD incidence in employees of a non-specific company. To parameterise this model, we first reviewed the evidence on the consequences this may have in terms of NCDs for teleworking employees, notably underlining potential impacts on mental health and MSDs. Our rapid review uncovered a wide variety of possible exposure-response relationships between teleworking intensity and NCD risk. By incorporating this data in our model, we showed that optimal frequency and timing of implementation of teleworking during an epidemic wave could vary widely. For instance, for health impacts associated with teleworking through a L-shaped function with a strength of association such as the one we identified for Edington risk (22), rapid and wide implementation of teleworking during the first few days of an epidemic can reduce both ID and NCD incidences. On the other hand, for a U-shaped relationship with parameters such as the one we identified for depression (26), intermediate (3-4 days per week) teleworking frequencies may be more optimal to maximise health benefits, while for inverted U-shaped relationships with parameters as observed for lower back pain (27), it may be necessary to weigh the increased NCD risk attributable to teleworking against the decreased ID risk. Importantly, both the shape and the upper/lower bound of these exposure-response functions must be taken into consideration when contrasting ID and NCD risk. We further demonstrate the need for this weighing by showing that telework can increase the relative importance of the health burden of NCDs, highlighting the importance of accounting for both ID and NCD risk when evaluating the impact of telework.

### 5.1. Implications

Through our rapid review, we confirmed that telework may impact both mental and physical health, although we identified several important knowledge gaps based on the selected studies. First, there is a lack of uniformity across the measurements of health outcomes (clinical diagnoses and declarative statements) explored in relation with teleworking, ranging from physical to mental health (depression, anxiety, burnout). Second, exposures are poorly characterised. Exposure to teleworking is not coded in a standardised manner; depending on the study, it may be expressed in terms of number of days per week, or number of hours per day, with different categories. Third, study contexts are heterogenous. Studies encompassing short- and long-term perspectives, within both pandemic and non-pandemic settings, yield outcomes potentially contingent on the phase of the COVID-19 pandemic (38). Confounding factors (such as the region, pre-existing comorbidities or professional status) are not well described or controlled for, and differ across studies. Fourth, very few studies met our inclusion criteria, and we found no longitudinal study that focused on MSDs, leading to a lack of quality in evidence especially regarding the temporal relationship between exposure and MSDs. Our findings are in agreement with two recent systematic reviews that also outlined the low quality of evidence regarding health impacts of teleworking (39,40). A recent study also shows that the relationship between telework and well-being can be dynamic over the duration of a pandemic and will differ according to the baseline level of telework (41).

Overall, the identified studies led to divergent results on the shape of the exposure-response function, depending on the considered health outcome (U-shaped, inverted U-shaped, L-shaped, broadly decreasing, and broadly increasing curves). In addition, even for the same health outcome, reported impacts of teleworking could be heterogeneous. For example, Henke and colleagues found a broadly decreasing relationship between hours/month of telecommuting and depression, while Chen and colleagues found a U-shaped relationship. This conflicting evidence may be due to modifying effects as demonstrated by one study according to which organisational factors within a company could alter the shape/direction of the relationship, depending on the COVID-19 wave (38).

Our mathematical model suggests that the shape and range of variation of the exposure-response relationship function between teleworking and NCD risk may influence the optimal teleworking frequency. When using this kind of model in an epidemic context, it is crucial to initially define the risk levels considered optimal for both ID and NCD. For a given target of incidence reduction, the choice of teleworking frequency and the timing of its implementation as an intervention during an epidemic wave varies. The implications in terms of performances and costs are also different as we expect sick-leaves due to IDs to be numerous but short while sick-leaves due to NCDs would be fewer but longer. Furthermore, MSDs and psychological disorders are both heterogeneous in severity, which makes quantitative assessments difficult. Finally, implications may largely depend on the professional sector, for example, workers in healthcare settings or densely-populated workplaces are expected to be more exposed to IDs, while desk-based workers will be more exposed to NCDs. The risk of ID and NCD, and therefore the definition of the “optimal” teleworking frequency could therefore vary depending on the job category and the associated tasks.

### 5.2. Limitations

As shown in our rapid review, the relationship between telework frequency and the risk of developing common NCDs such as lower back pain or psychological distress in the long-term depends on many unmeasured individual and environmental characteristics. Instead of accounting for all these specificities, we illustrated with our model the impact of different relationships considering one average risk for all individuals. The lack of significant associations between telework and some NCDs is also a source of uncertainty in the exposure-response relationships used in the model, as shown in Table 2. Alternative modelling strategies could be used to integrate individual heterogeneity, but this would require additional evidence regarding the distribution of exposures and risks based on individual characteristics (age, gender, job…) that is not yet available in the literature. In addition, one of our associations (inverted U-shaped relationship for lower back pain) is derived from a single cross-sectional study, limiting causal inference and highlighting the scarcity of evidence on teleworking and NCD incidence. Another key limitation of the current evidence base linking telework and NCDs is its reliance on arbitrarily defined categories of telework exposure, as no study to date has reported fully disaggregated or continuous measures. This may bias the estimation and shape of dose–response relationships, and affect the conclusions of studies like ours.

The majority of evidence regarding telework arises from the COVID-19 pandemic. In this context, telework was frequently unplanned and imposed on individuals, which does not necessarily reflect conditions where teleworking would be planned and adapted at an individual level (7). In addition, relatively few countries were represented in these studies, while we would expect the impact of teleworking to vary across regions, between urban and rural settings and living conditions. Finally, the impact of teleworking on health likely depends on the socio-professional categories considered and its desirability.

In our illustrative example, we simulated the impact of telework policies in the context of the second wave of the SARS-CoV-2 pandemic. Thus, the results should be interpreted in the context of an emerging ID for which no pharmaceutical intervention (e.g., vaccination) is available, and for which no behavioural change among workers is observed apart from the telecommuting policy decided by the employer. We expect different results in case these two assumptions are not met.

Finally, we assumed homogeneous mixing within the company, whereby all employees could be in contact, without considering more complex work organisations. Similarly, we considered a simple telework policy according to which a fixed percentage of the total workforce in the company is teleworking every day, but more refined policies have been implemented during the pandemic, such as rotating telework (13). In these strategies, employees are evenly distributed in groups that alternate on a daily or weekly basis. This is expected to reduce the overall number of contacts per individual and potentially the risk of transmission, whilst maintaining a reduced average frequency of telework. Building a discrete-time model to represent alternating teleworking days and on-site days would be a valuable extension, providing a more realistic description of telework patterns and transmission dynamics.

### 5.3. Perspective and future directions

Several options could be considered to mitigate the NCD risk among teleworking individuals. First, sedentary behaviours, prolonged computer use, and poor ergonomics during telework have been linked to increased musculoskeletal disorders (MSDs), particularly affecting the neck, shoulders, and lower back. To mitigate MSD risks, studies recommend ergonomic workspaces, regular posture changes, and proactive telework preparations by companies, ensuring adequate support and proper work conditions. Second, telework has mixed effects on mental health, enhancing positive emotions, job satisfaction, and organizational commitment while reducing emotional exhaustion, but also contributing to anxiety, stress, fatigue, and social isolation. Negative effects could be mitigated by measures like technical support, flexible hours, social communication, and health interventions, with part-time telework offering benefits for work-life balance and social relationships.

In practice, the optimal frequency and implementation timing of telework will depend on the relative importance given to both NCDs and ID, as we show by estimating the health burden of both diseases under varying teleworking frequencies. The decision on respective weights given to the ID and NCD incidence should consider the different timelines at which these diseases occur: typically, short-term for IDs and mid- to long-term for NCDs. IDs represent a substantial socioeconomic burden (42), which can justify the implementation of teleworking as an intervention to reduce disease spread. Part of this burden is related to sick leave (43), which can lead companies to act in order to minimise disease incidence among their employees. However, when designing a teleworking policy, deciders need to account for feasibility, legal and ethical criteria. For instance, teleworkers need to have access to telework equipment, which was not always straightforward during the COVID-19 pandemic (44).

Future studies should better characterize the distribution of telework among population and identify its association with NCDs and mental health outcomes based on individual characteristics (age, gender, job…). It is also important to explore how telework dynamics can influence the transmission of infectious diseases among workers.

Current lack of evidence, especially data to assess the impact of exposure to telework more finely, prevented us from relying on robust exposure-response functions linking NCDs and telework. However, pending such data, our work provides a framework to balance the infectious and NCD risks in an epidemiologic context that could easily be re-used with updated data.

## 6. Conclusions

In our rapid review, we identified three studies, two being longitudinal. This very low number underlines the need for more data to monitor the health impacts of teleworking. In particular, further studies should characterise the relationship between telework frequency and NCDs. To this end, it is crucial to collect data outside the context of the COVID-19 pandemic, since additional stressors during this period may have modulated the relationship between telework and health. In addition, the mechanisms by which teleworking impacts mental and physical health should be better characterised (e.g. unadapted workstation for MSDs). Lastly, further efforts are needed to identify the individual factors affecting the exposure-response relationships both during and outside of epidemic contexts.

Our innovative approach, which attempts to combine short- and longer-term consequences of teleworking in a unique framework, could serve as a basis to develop tools for employers and policymakers. Such tools could be used to quantify the impact of telework on employee health and identify optimal telework strategies to limit health adverse events and improve employee well-being.

## Data Availability

All code used in the analysis are available in the following GitHub repository:

https://github.com/MESuRS-Lab/telework_health

## 8. List of abbreviations

ID: Infectious disease
IU: Inverted U-shaped
LD: Linear decrease
LI: Linear increase
LS: L-shaped
MSD: Musculoskeletal disorder
NCD: Non-communicable disease
US: U-shaped

## 9. Declarations

### Ethics approval and consent to participate

Not applicable.

### Consent for publication

Not applicable.

### Data, script and code availability

All code used in the analysis are available in the following GitHub repository: https://github.com/MESuRS-Lab/telework_health. The code at the time of publication is archived online at: https://doi.org/10.5281/zenodo.19738028.

### Supplementary Information

The Supplementary Information for this article can be accessed online (https://doi.org/10.5281/zenodo.19738147). This file includes “Supplementary Text 1:

Calculation of the basic reproduction number R0”, and “Supplementary Text 2: Calculation of the exposure-response functions between teleworking and non-communicable disease risk” (including Supplementary Figures 1-4).

### Competing interests

The authors declare that they have no competing interests.

### Funding

This project did not receive any specific funding.

### Authors’ contributions

All authors contributed to the design of this study. LM, FT, EH, NL, KAB, and MBH performed the study selection for the rapid review based on our inclusion and exclusion criteria. QJL, ML, PH, AM and LT designed and implemented the mathematical model. WB, SZ and KJ contributed to data analysis. All authors contributed to the redaction of the manuscript. All authors read and approved the final manuscript.

## Acknowledgements

The authors would like to thank Hanifa Bouziri for helpful discussions on musculoskeletal disorders, and Pascal Crépey for access to the COVID-19 incidence data.

## Notes

### Competing Interest Statement

The authors have declared no competing interest.

### Summary of Updates

Peer-reviewed version recommended by PCI

